# Barriers to Cochlear Implant Uptake in Adults: A Scoping Review

**DOI:** 10.1101/2024.05.15.24307334

**Authors:** Jonathan D. Neukam, Ansley J. Kunnath, Ankita Patro, René H. Gifford, David S. Haynes, Aaron C. Moberly, Terrin N. Tamati

## Abstract

**Introduction:** Cochlear Implants (CIs) provide access to sound and help mitigate the negative effects of hearing loss. As a field, we are successfully implanting more adults with greater amounts of residual hearing than ever before. Despite this, utilization remains low, which is thought to arise from barriers that are both intrinsic and extrinsic. A considerable body of literature has been published in the last five years on barriers to adult CI uptake, and understanding these barriers is critical to improving access and utilization. This scoping review aims to summarize the existing literature and provide a guide to understanding barriers to adult CI uptake.

**Methods:** Inclusion criteria were limited to peer-reviewed articles involving adults, written in English, and accessible with a university library subscription. A cutoff of 20 years was used to limit the search. Barriers uncovered in this review were categorized into an ecological framework.

**Results:** The initial search revealed 2,315 items after duplicates were removed. One hundred thirty-one articles were reviewed under full-text, and 68 articles met inclusion criteria.

**Discussion:** Race, ethnicity, and reimbursement are policy and structural barriers. Public awareness and education are societal barriers. Referral and geographical challenges are forms of organizational barriers. Living context and professional support are interpersonal barriers. At the individual level sound quality, uncertainty of outcome, surgery, loss of residual hearing, and irreversibility are all barriers to CI uptake. By organizing barriers into an ecological framework, targeted interventions can be used to overcome such barriers.

## Introduction

Two out of three adults over the age of 65 have hearing loss, which may be associated with impaired cognitive function, decreased physical activity, and poor health care utilization (1–3). Cochlear implants (CIs) provide access to sound, help mitigate the negative effects of hearing loss, and improve communication ability. Traditionally, implantation was limited to adults with bilateral profound sensorineural hearing loss; however, the degrees and configuration of hearing loss required for implantation have become less stringent. As a field, we are successfully implanting more adults with greater amounts of residual hearing, including those with asymmetric hearing loss and single-sided deafness. Despite the expansion of candidacy criteria, utilization rates of CIs remain low. Only between 2% and 13% of eligible adults in the United States (US) receive a CI (4,5). Additionally, demographic disparities exist in those who pursue cochlear implantation, with minorities making up an even smaller percentage of those being implanted compared to local or regional demographics (6–9).

Although a considerable body of literature has been published on barriers to hearing aid (HA) uptake (for recent review see Knoetze, et al. (10)), our understanding of the barriers to CI pursuit is still limited. While some barriers to HA and CI use may overlap, cochlear implantation presents unique challenges, such as lack of insurance reimbursement or out-of-pocket expense for a high-cost device and surgery, surgical considerations, and potentially greater uncertainty of outcome for an irreversible intervention. Better defining and understanding these barriers can help clinicians address questions and concerns from patients who can make better-informed decisions about their hearing healthcare and, in turn, improve CI utilization rates, especially among underserved populations. The need for this understanding is further emphasized by the limited number of comprehensive, global reviews written on adult CI uptake barriers. A thorough review of adult and pediatric barriers in the US was written over 10 years ago, followed by a few opinion articles in recent years (11–14), highlighting a clear need for an updated guide for understanding global barriers in adult CI uptake.

This scoping review aims to summarize the existing literature on barriers to pursuing CIs in adults. Preliminary evidence suggests that barriers to CI uptake can be compared to broad barriers in health behavior, which can be organized into an ecological framework with overarching barriers at the system and policy levels down to the individual level. Models of health behavior are not novel to public health. These models can aid in identifying targeted interventions to advance health utilization (15–17). Therefore, the focus of this review is to provide a guide regarding (1) the types of studies completed to this date, (2) the types of materials and individual characteristics that can impact barriers to CI uptake, and (3) the gaps in our knowledge by providing a framework with suggestions for future research.

## Methods

The Preferred Reporting Items for Systematic Reviews and Meta-Analysis extension for Scoping Reviews (PRISMA-ScR) was used to guide reporting. Inclusion criteria were limited to peer-reviewed articles involving adults, written in English, and accessible with a university library subscription. A cutoff of 20 years was used to limit the search (2003–2023) because, during that time range, CIs have been widely viewed as the standard of care for adults with moderate-to-profound sensorineural hearing loss (18). A preliminary review was conducted by the first author. Twenty-nine articles were collected and reviewed, which expanded the ability to search with more inclusive terms. Any study (prospective or retrospective) that gave insight into factors related to CI uptake and barriers was included. Opinion and review articles were also included. This review study was exempt from institutional Review Board approval.

### Search Strategy and Extraction

A search string was designed using the National Library of Medicine’s Medical Subject Heading (MeSH) browser. Title and abstract MeSH terms associated with “cochlear implant” and “barriers,” including disparities, socioeconomic, and vulnerable populations, were used. The search was performed on PubMed and the search string was translated for Embase, CINAHL, PsycINFO, Web of Science, Cochrane, and ERIC. The search was performed in September 2023. See supplementary material for search string. All items were imported into Covidence (Veritas Health, Melbourne, Australia) and screened independently by two reviewers. A third reviewer was involved if an item did not meet eligibility criteria by both reviewers one and two.

### Quality and Risk of Bias Assessment

The level of evidence for each article was assessed using the Oxford Centre for Evidence-based Medicine guidelines (19). Based on the review type of reporting qualitative and quantitative research with no planned approach of a systematic review, we determined *a priori* not to formally assess risk of bias.

## Results

The initial search revealed 4,127 items, with 2,315 remaining after duplicates were removed. Titles and abstracts were reviewed and yielded 131 articles for full-text review. 67 articles met inclusion criteria. One additional article published shortly after the search date was identified by the authors and included due to its relevancy. See Figure 1 for schematic.

**Figure 1:**
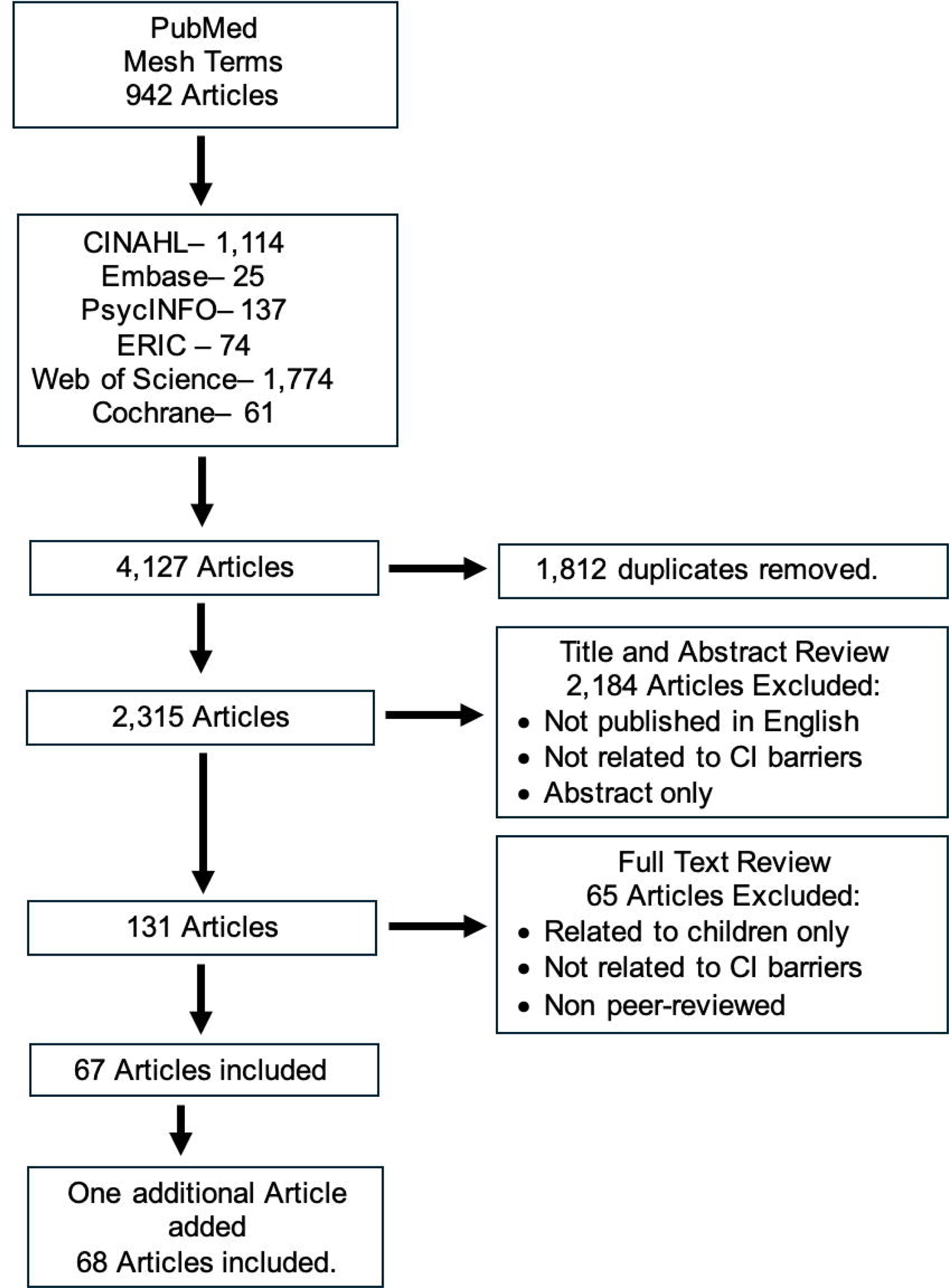
Schematic of Search.

The majority (69%) of the articles have been published since 2020 (Figure 2). Most articles focused on barriers in the United States (US) (46%), followed in frequency by the United Kingdom (UK) (16%). Fifty-six studies were comprised of 28 (41%) retrospective studies, 27 (40%) prospective studies. Three (4%) were combined type studies. There were six opinion articles, one summarized interview panel, and three review articles related to general barriers in hearing healthcare, reporting of sociodemographic data in CI clinical trials, and CI delivery models. See Table 1 for a list of articles and characteristics, including level of evidence.

**Figure 2:**
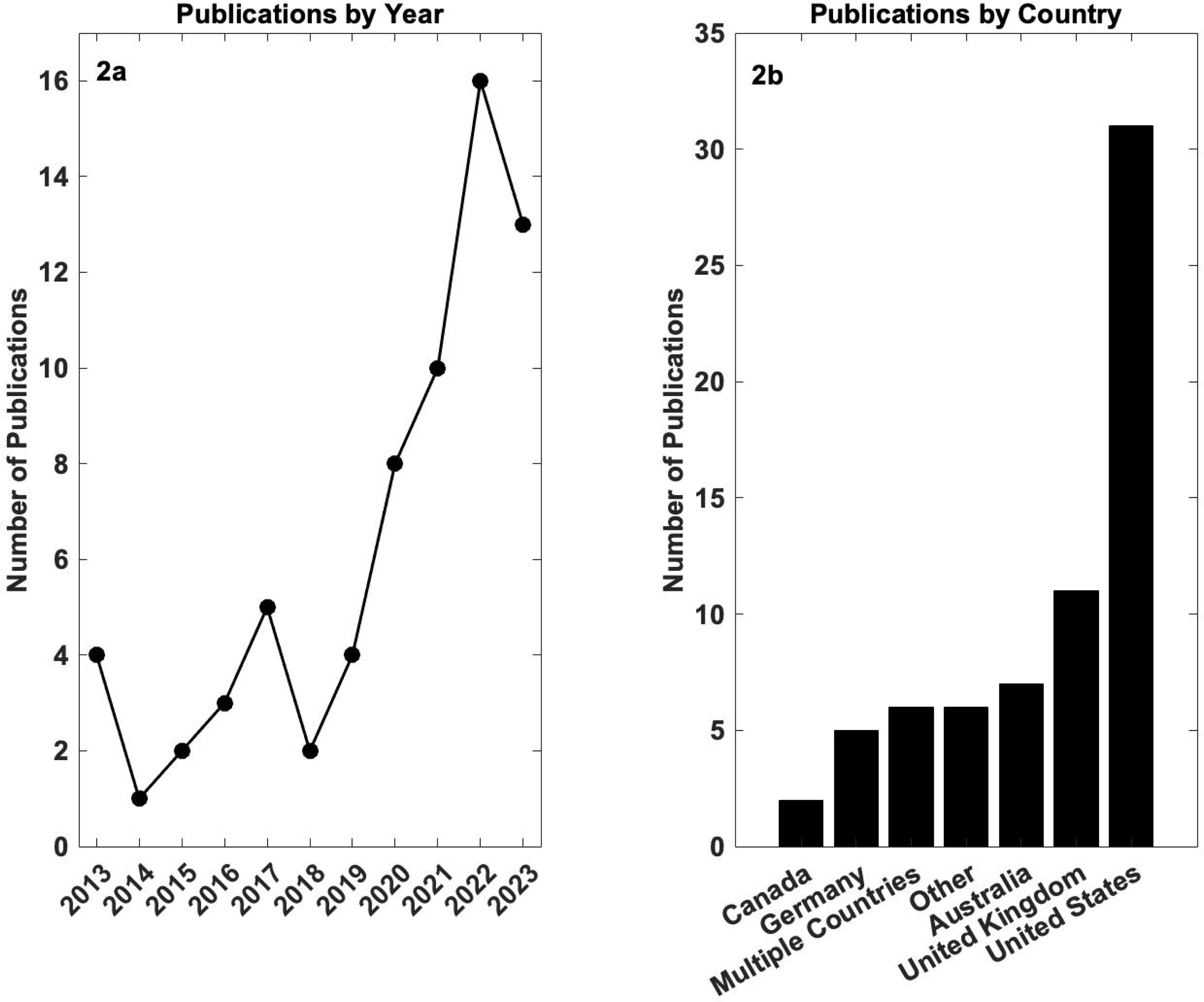

**Table 1:**
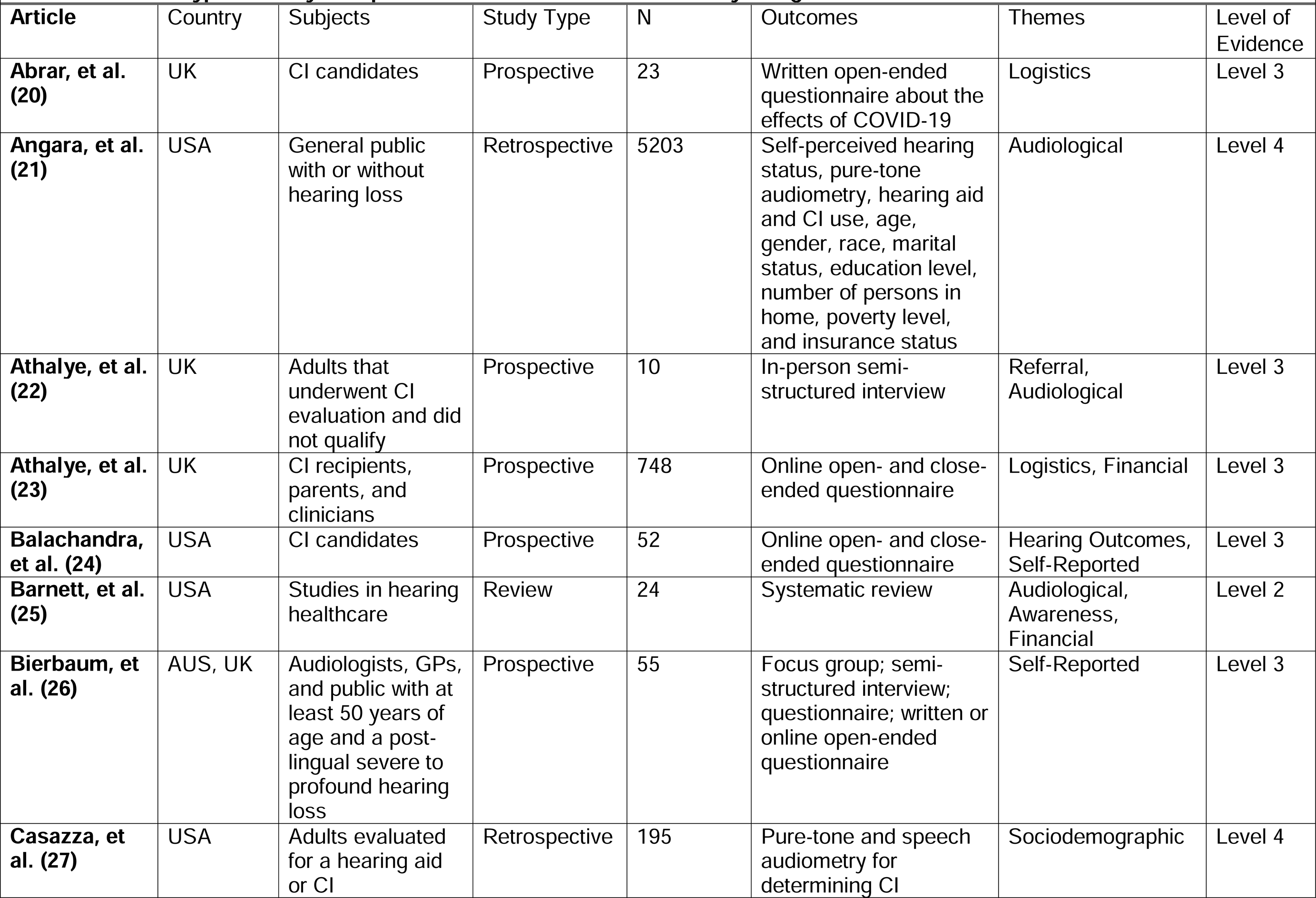
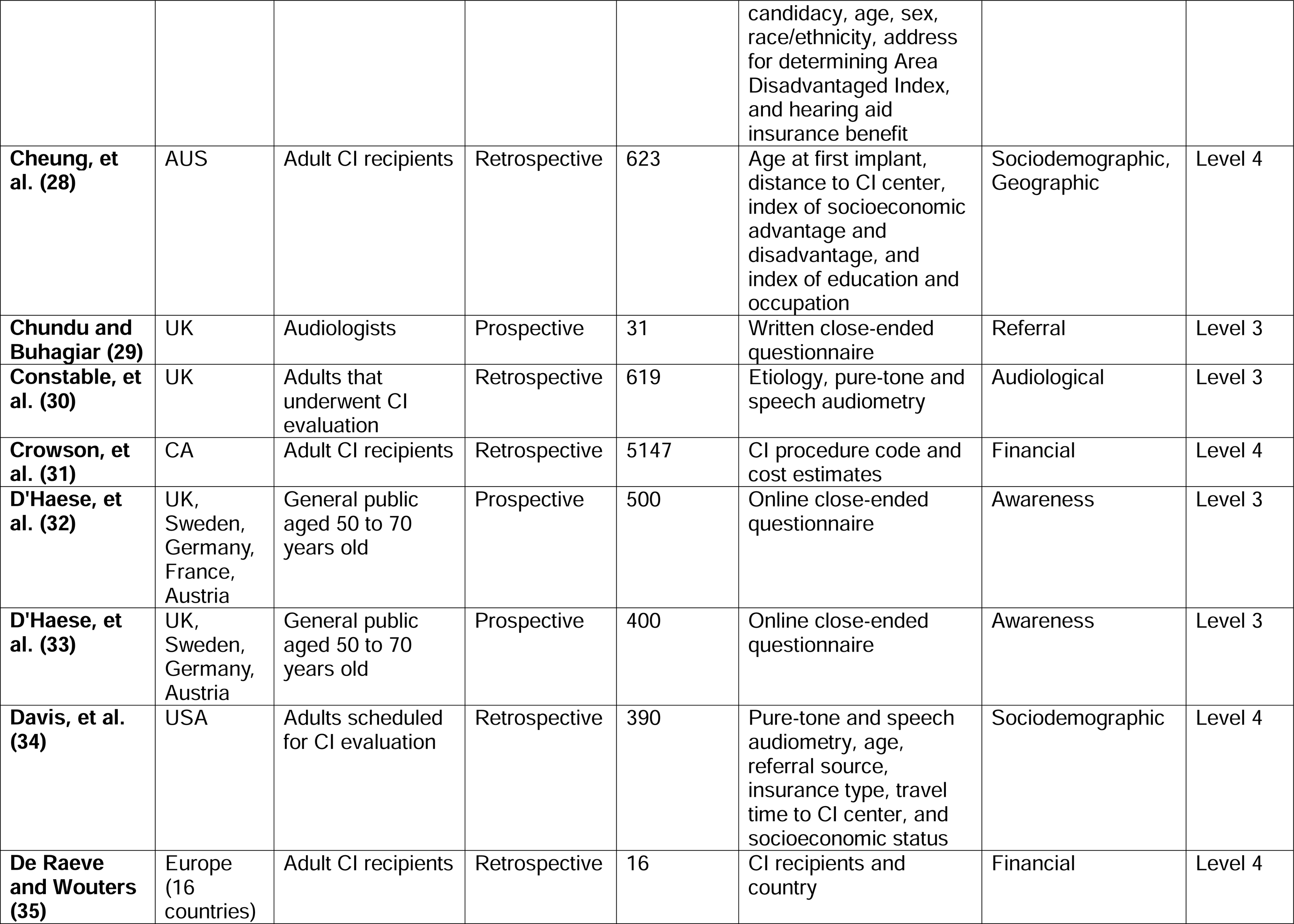
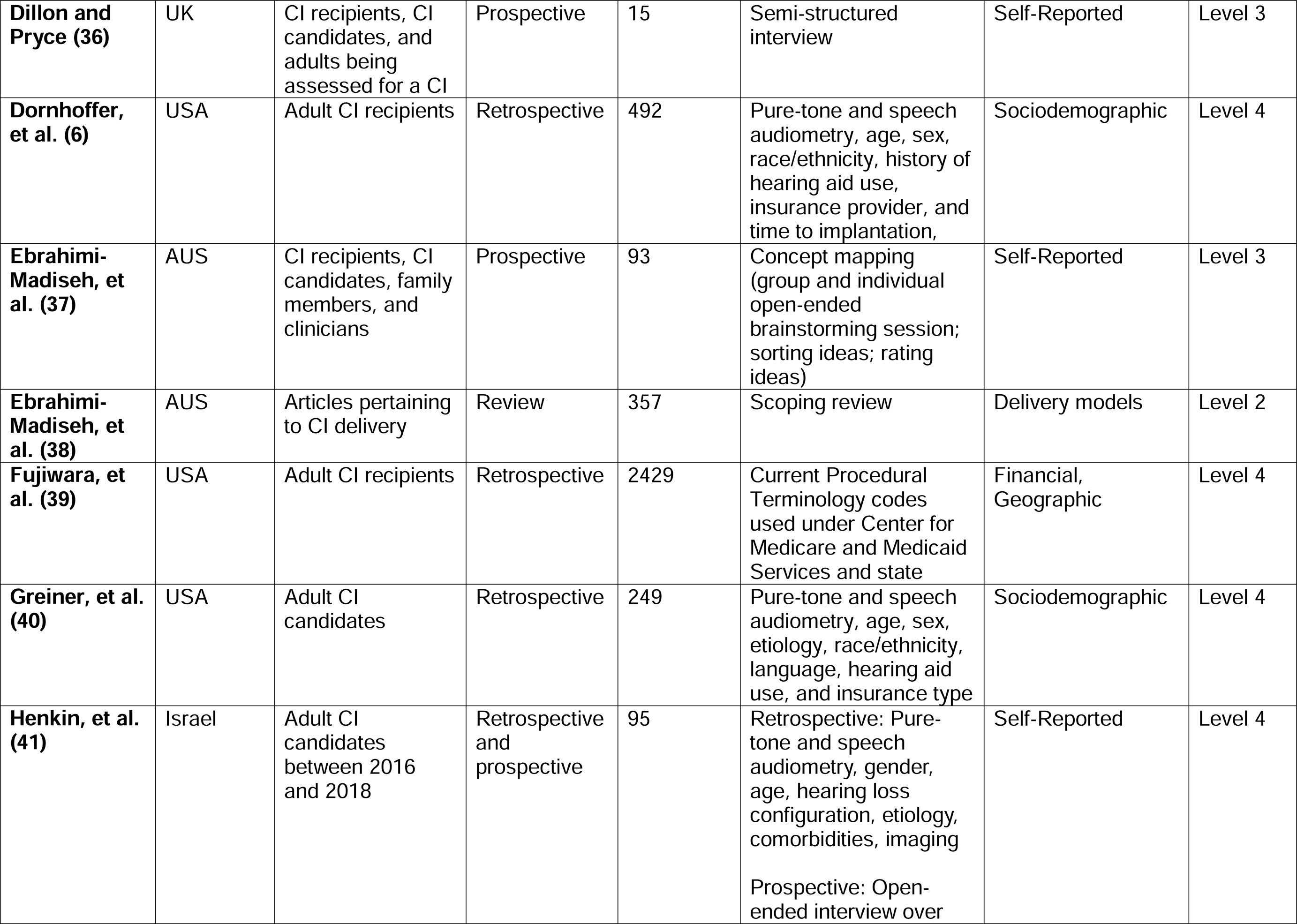
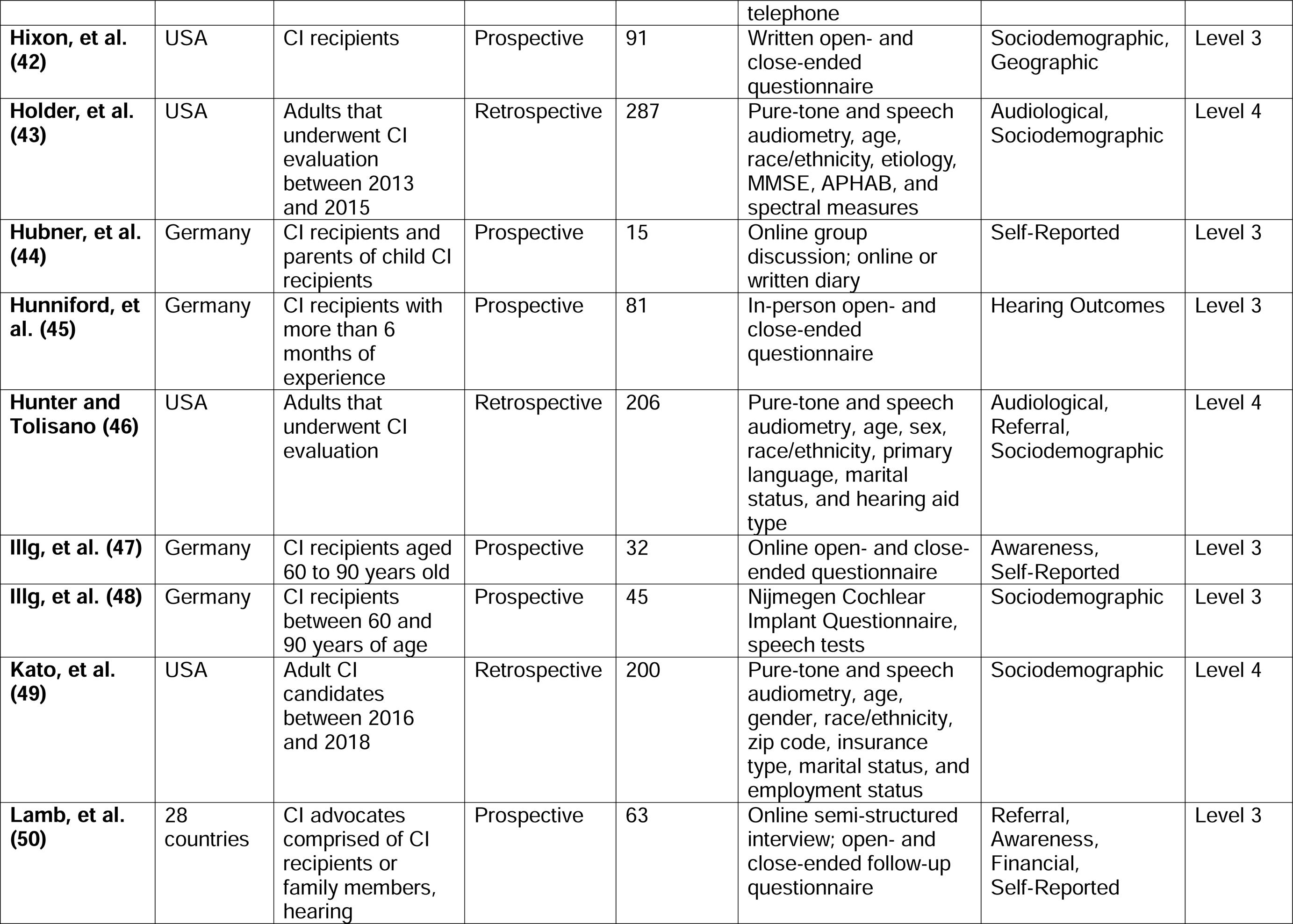
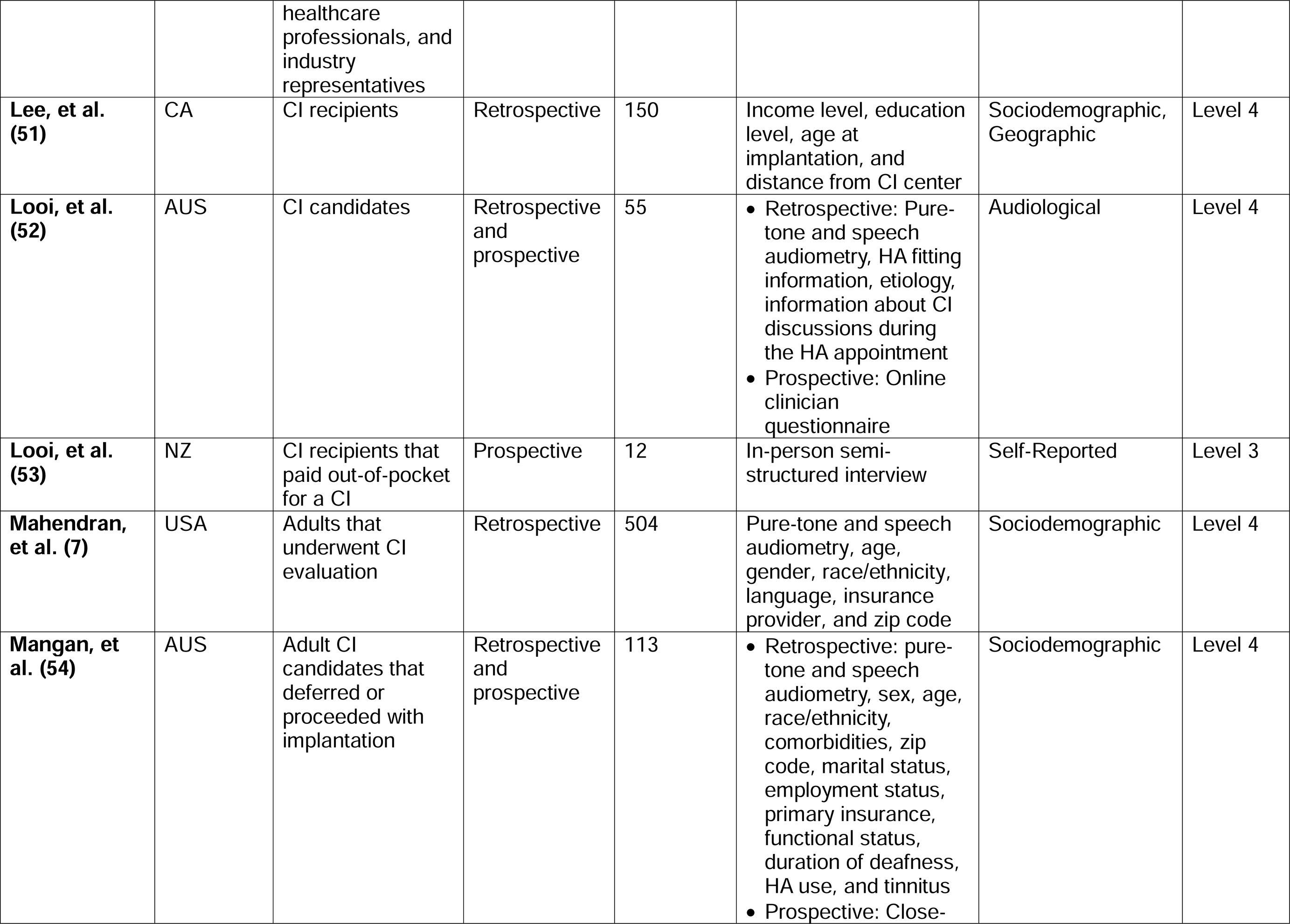
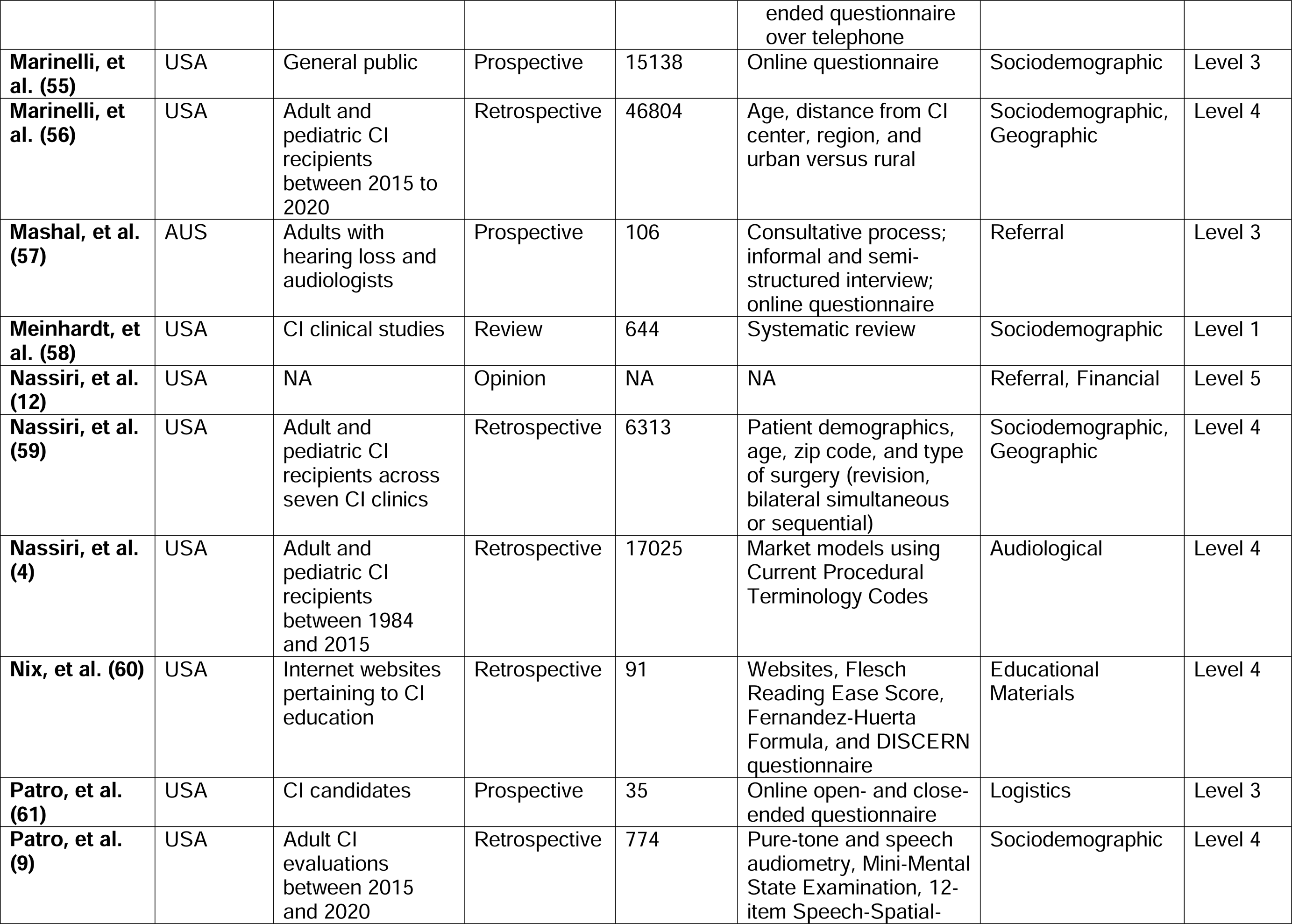
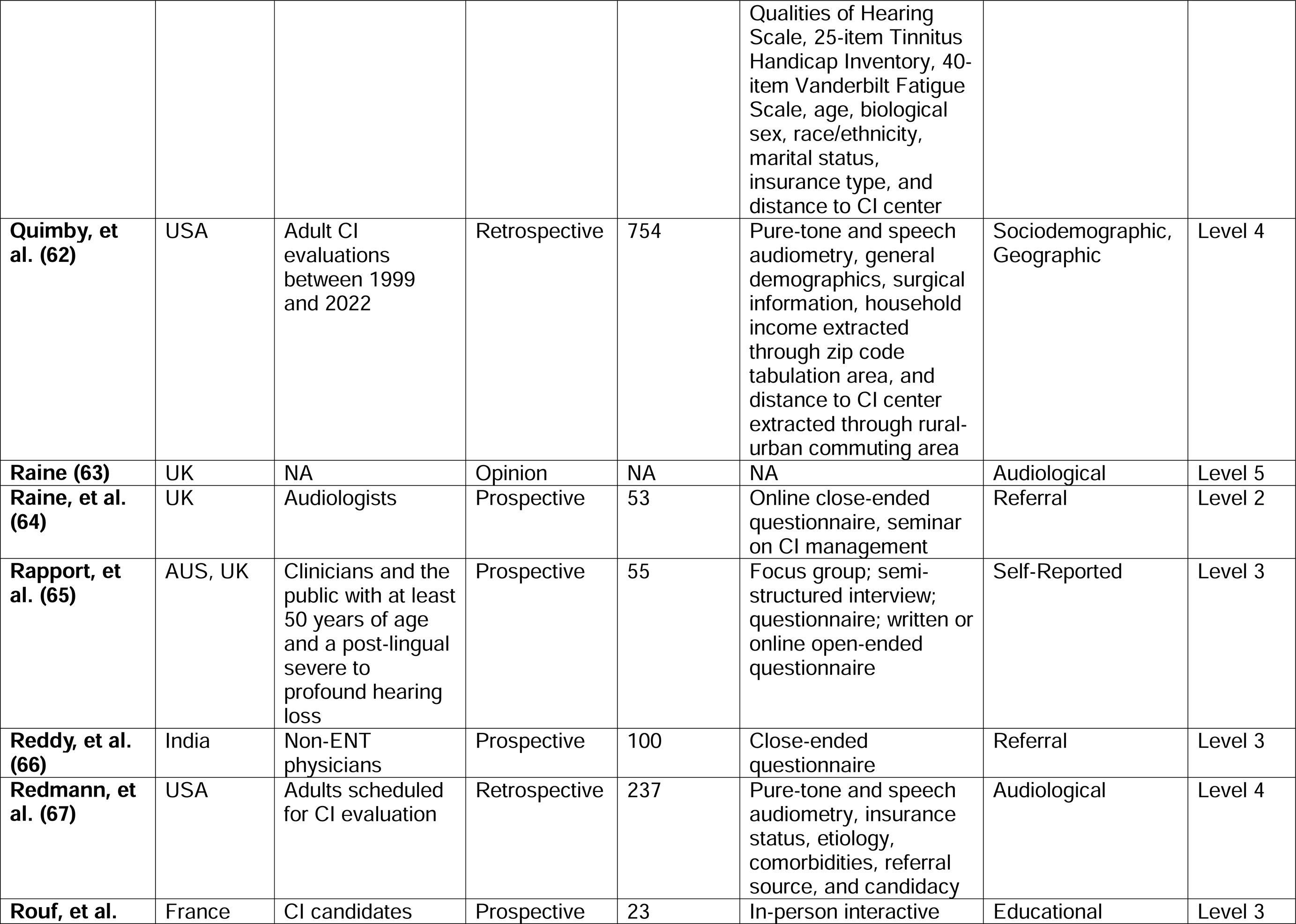
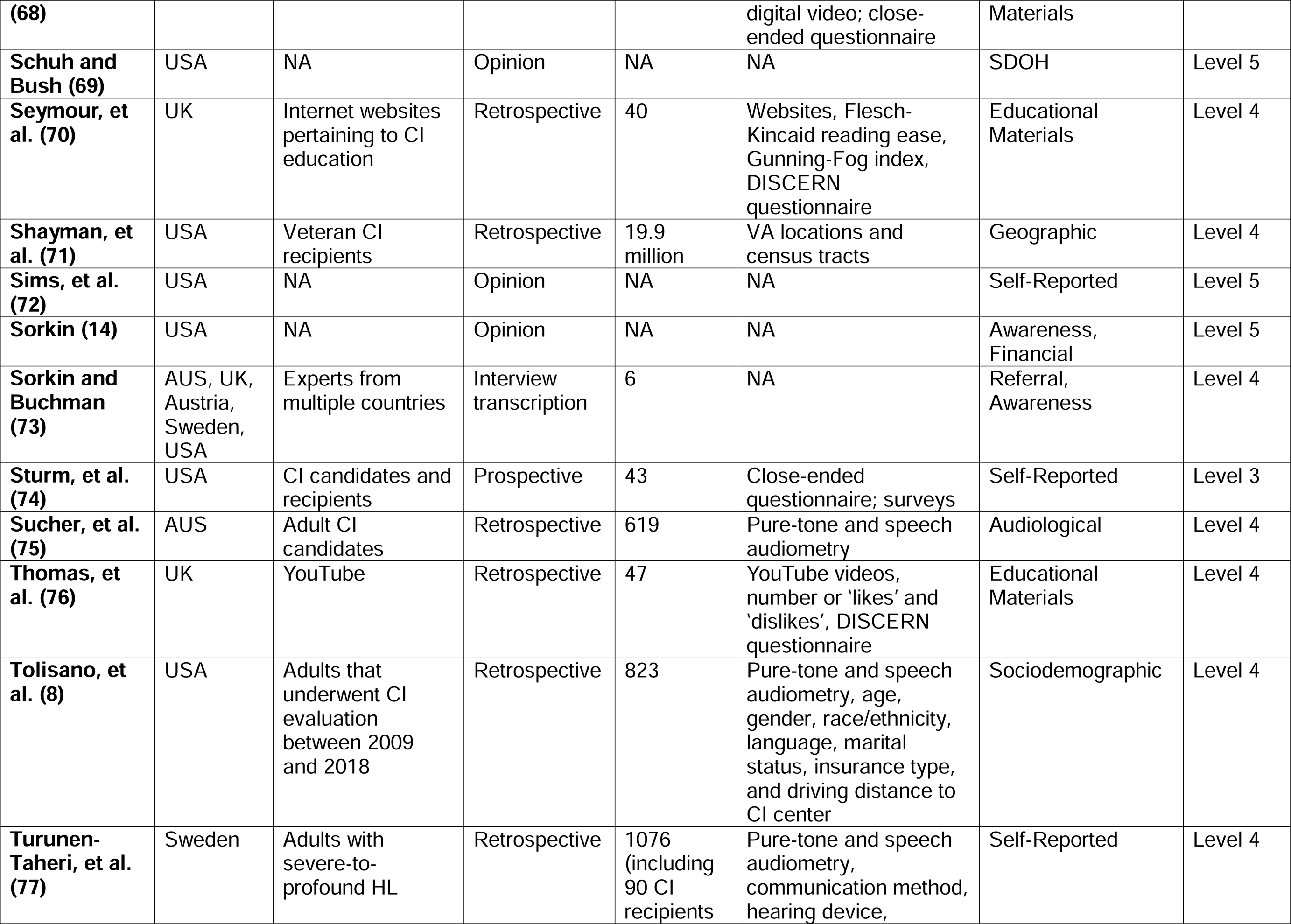
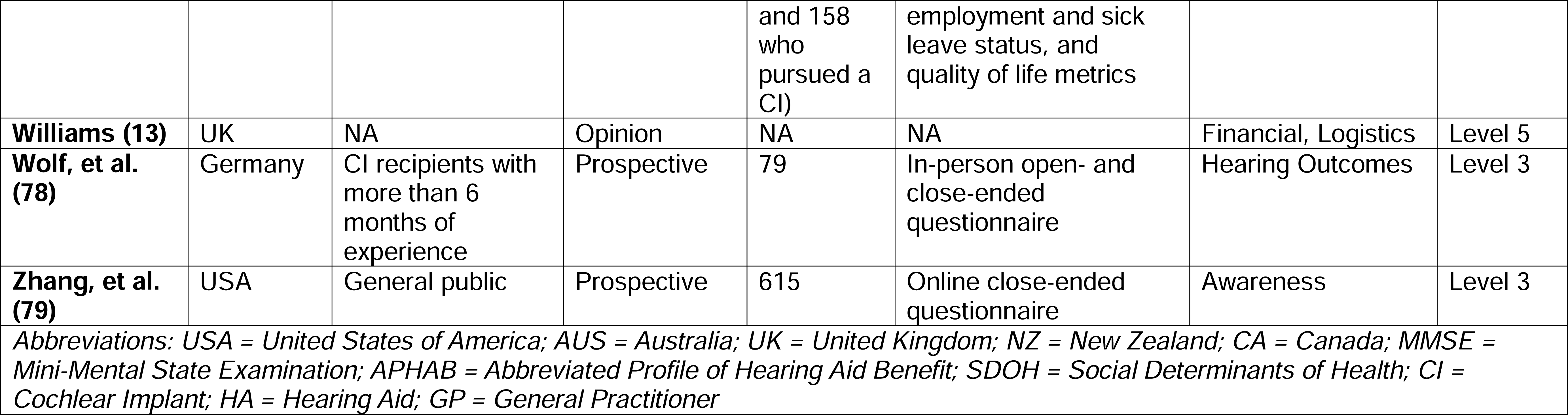
List of studies and articles included in review with country of publication or study, characteristics of the population or material studied, type of study, sample size, tool utilized or factors analyzed, general themes, and level of evidence.

### Policy and Structural

A summary of barriers can be seen in Table 2 organized into a five-category ecological framework. Fifteen retrospective studies (6–9,27,28,30,31,34,35,39,49,52,59,62) and five prospective studies (20,22,23,42,43) examined policy and structural factors that impact CI pursuance. In the US, several studies showed that non-Caucasians pursued CIs at a lower rate but qualified for CIs at higher rates (6–9,27,49). In multiple countries, socioeconomic status and education level were found to be factors in CI uptake (28,34,51,62). Eligible adults with higher incomes and higher education levels were more likely to pursue a CI. In countries with single payer or universal healthcare, financial budgets may restrict the number of CIs to be performed, and country-specific eligibility criteria can also inhibit those seeking a CI (22,23,30,31,35,39,73).

**Table 2:** Summary of barriers with citations formatted into an ecological framework.

| **Policy and Structural Barriers** |
| --- |
| • Structural racism (6-9,21,27,49,69)<br>• Lack of health insurance/ inadequate coverage (9,21,39,40,69)<br>• Socioeconomic status (21,28,34,62,69)<br>• Education level (21,51)<br>• Rural/urban status (56,59,62,69)<br>• Budget policies restricting the number of CIs (23,31,35,69)<br>• Country-specific eligibility criteria (4,22,30,31,46,50,69)<br>• Global pandemic (20,56,61) |
| **Societal Barriers** |
| • Lack of awareness about CIs by public (20,32,44,50,55,79)<br>• Misconceptions about CIs by public (20,32,44,55,79,80)<br>• Education materials (20,60,68-70,72,76,80)<br>• Decreased reliance on professionals for education (33)<br>• Stigma of hearing loss (36,72,80) |
| **Organizational Barriers** |
| • Lack of skills by professionals (22,23,26,29,80)<br>• Lack of awareness from professionals (20,29,50,52,57,64,66)<br>• Inappropriate tests for referral (40,50,57)<br>• Inconsistent referral criteria (20,26,50,52,53,57,59)<br>• Geographic barriers of CI centers and audiology clinics (26,28,42,51,53,56,59,71) |
| **Interpersonal Barriers** |
| • Lack of social and professional support (20,22,26,50,69)<br>• Living context/marital status (9,20,36,54)<br>• Lack of community among CI users and candidates (26,50) |
| **Individual Barriers** |
| • Uncertainty of outcome (20,24,26,37,47,53)<br>• Age (6,21,49)<br>• Fear of surgery (9,43,74,79,81)<br>• Sufficient HA performance (20,67,77,81)<br>• Hearing loss configuration (82)<br>• Inconsistency between perceived and measured hearing deficit (21,74)<br>• Financial (20,26,53,54,67,69,72,74) |
| *Abbreviations: CI = Cochlear Implant; HA = Hearing Aid* |

Although rare, another example of a policy and structural barrier is a global pandemic (Coronavirus-19). An analysis from two CI manufacturers in the US showed up to a 25% reduction in adult surgeries during the height of the coronavirus pandemic in 2020 (56). Many hospitals temporarily halted elective surgeries to redirect hospital staff and resources and to prevent unnecessary infections. A qualitative study out of the UK found that patients who had their CI surgery delayed by no more than three months due to the pandemic were disappointed and reported that the delay negatively impacted their mental health (20).

### Societal

Three retrospective studies (60,70,76) and ten prospective studies (26,32,33,44,47,50,53,55,68,79) examined societal barriers. Inaccurate perceptions about CIs and lack of awareness are barriers to uptake. When probing the public’s general knowledge of CIs, it was found that many believed that CIs were fully implantable without the need for an external speech processor (32) and that the internal components must be replaced every three to five years (79). These misconceptions of cochlear implantation may stem from reliance on the internet rather than healthcare professionals for knowledge about CIs (32,33,47). Moreover, internet sites providing CI education were found to be written higher than a sixth-grade reading level, which is the recommended reading level by the American Medical Association (60,70). Educational materials such as YouTube videos on CIs often did not have subtitles or interpreter services (76). Only 12% of the general population was aware of this technology, with greater awareness among young adults and Caucasians (55).

### Organizational

Inconsistent referral criteria and candidate identification as well as lack of professional resources are examples of barriers at the organizational level. Lack of professional resources can be due to geographical challenges such as a limited number of CI centers in a region, or centers with audiologists that are not properly trained and competent in CI programming. CIs require follow-up programming appointments and sometimes aural rehabilitation. Three retrospective studies (40,46,71) and 12 prospective studies (22,23,26,29,44,52,53,57,61,64–66) examined barriers at the organizational level. Inconsistent referral criteria were reported by audiologists, and lack of awareness among non-audiology professionals was also reported (26,29,44,50,57,64–66). Insufficient testing materials, such as speech recognition materials that are in a patient’s primary language, can result in the inability to test aided-speech performance. This can lead to further delays in the CI referral process, especially in minority populations (40). Geographically, patients in rural areas travel farther to appointments because many CI centers are located in urban areas (42,51,56,59,71).

### Interpersonal and Individual

At the patient level, barriers can exist that are both interpersonal and individual. A single retrospective study (54) and three prospective studies (26,50,72) examined interpersonal barriers, while six retrospective studies (21,51,61,67,77,82) and 13 prospective studies (24,26,36,37,43,47,48,53,54,72,74,78,81) examined individual barriers. Marital status and living context (i.e., living alone versus cohabitation) were shown to be predictors of CI uptake, suggesting that the lack of strong support systems can be a barrier (9,36,49,54). Furthermore, establishing a network of existing CI recipients to answer questions and share their experiences can positively impact CI pursuit. Potential candidates have benefitted from meeting other CI users (26,50). Individual barriers to CI uptake are strongly tied to a patient’s values, beliefs, and fears. Sound quality, uncertainty of outcome, surgery, the loss of residual hearing, irreversibility, and a general sense of not being ready for a CI were the top-rated individual barriers by CI recipients (24,26). Self-perceived hearing loss and hearing difficulty, the social stigma of hearing loss or the use of hearing-related technology, and aesthetics can also be barriers to CI uptake (53,72,74). Age has also been found to play a negative role in the decision to pursue a CI (6,49,77).

## Discussion

The intent of this review was to shed light on barriers to CI uptake in adults. The field of cochlear implantation has evolved greatly in the last two decades. Some examples are the FDA approval of CIs for single-sided deafness, implementation of electroacoustic stimulation for preserved low-frequency hearing, remote programming options, MRI compatible internal retention magnets, and newly adopted eligibility criteria by the Center for Medicare and Medicaid Services in the US (83). These changes have broadened the use of CIs in adults. Patients are being implanted sooner compared to previous years, and some early obstacles have been overcome with advancements in technology. Despite the advancements, many adults have remaining trepidation and wait an average of 24-30 years with hearing loss before implantation, with longer wait times for patients in rural settings (6,42). Longer durations of deafness have been associated with worse speech recognition outcomes after cochlear implantation (84–86).

In an opinion paper over ten years ago, Sorkin 2013 (14) identified the main barriers to CI uptake in the US – CI awareness, referral, reimbursement, best practice guidelines, data on cost effectiveness, and political issues surrounding deafness. Since 2013, best practices have evolved substantially. Data on cost-effectiveness has partially driven the increase in CI utilization and reimbursement by showing that unilateral implantation leads to sustained changes in quality of life across the lifespan (for review see Crowson, et al. (87)). Political issues surrounding deafness have also lessened. In 2018, the American Speech-Language-Hearing Association issued a position statement that recognized American Sign Language as a unique language (88). Additionally, Gallaudet University, a US-based culturally Deaf university, developed the Bilingual Mission Framework in 2019, which promotes a language-rich environment through the use of ASL in conjunction with spoken English.

A key takeaway from this review is that race/ethnicity presents a structural barrier to CI uptake. This barrier is gaining increased visibility in the field by emerging research in the form of retrospective studies. Racial and ethnic disparities in CI use have previously been documented in children and are especially relevant to private payer systems like the US, where health disparities exist due to insurance coverage that can be linked to societal position (e.g., 28,89,90). Many retrospective studies in the US have uncovered that individual patient cohorts do not represent the racial and ethnic distribution of their region (6–9,27,43,49). As a field, we are disproportionately implanting more Caucasians and those from higher SES than any other groups.

Minority populations are underserved in many domains of healthcare, including the provision of HAs and CIs (91). This is likely a complex problem that may be related to community healthcare access and SES, which may be intertwined with race/ethnicity. Fully understanding the individual impact of each barrier and how these barriers interact with healthcare access is difficult. Countries with universal healthcare, such as Australia, still find disparities amongst SES groups, suggesting that this barrier extends beyond insurance coverage and may include factors such as the cost of travel to CI centers or the impact of surgical recovery and rehabilitation on one’s occupation (28). Sociodemographic underrepresentation creates biases in CI clinical research, impeding the generalization of rehabilitation methods or outcomes to the general population. This fact warrants more research and investigation into barriers for minority populations and patients from low SES backgrounds.

A lack of referral from hearing healthcare and non-hearing healthcare professionals can be one of the largest barriers to CI uptake. This issue may arise from HA audiologists not appropriately referring patients for a CI evaluation when a HA no longer provides functional benefit (92). One challenge is that functional or aided word recognition testing is not a standard audiological test performed outside of a CI evaluation. Some audiology clinics lack booth space or equipment for aided testing. Even if aided testing is performed, some audiologists are not trained on the current expanded eligibility criteria.

Lack of referral may also arise from other healthcare professionals. For instance, most appointments with general practitioners take place face-to-face in a quiet room with visual cues, which can limit awareness of the severity of hearing loss (12). They may not screen for hearing loss or communication issues as part of routine care, and there is insufficient evidence that hearing screening is beneficial in asymptomatic adults (93). However, self-reporting of hearing loss is not a consistent indicator of actual hearing deficit (21). Additionally, since CIs were originally intended for bilateral severe to profound hearing loss, some healthcare professionals may be unaware of expanded eligibility or skeptical about their ability to improve hearing in patients with less severe degreess of hearing loss.

Considering all barriers to CI pursuit that were identified in this review, geographic barriers were not universally found across countries. Distance and limited access to CI centers were relevant to CI uptake in Australia, New Zealand, UK, and Canada (26,51,53). In the US, patients who reside in rural areas travel longer distances to CI centers, but distance to CI center was not found to be a barrier (56,59,62). This could be due to differences in healthcare systems. Single payer systems might be implanting patients from a more economically heterogenous population. Private payer insurance coverage in the US is associated with economic stability, and, therefore, travel resources may not be an economic burden. The recent rollout of remote programming options by some CI manufacturers may help to alleviate some of the travel burden patients may encounter. Some CI centers are optimizing patient travel time with same-day surgeries (61,94), and some are overcoming fear of surgery by the use of local anesthetic (95,96). These advancements may also foster avenues to bring CI technology to third-world nations where very little is known about CI uptake. This is a major gap in our knowledge base of CI utilization and missing from this scoping review.

### Future Directions

With a comprehensive understanding and framework of barriers that exist, targeted interventions can help to overcome such barriers. Audiology and hearing healthcare is a rapidly changing field with new technologies and therapies being developed to meet the needs of our patients. The recent rise of over the counter (OTC) HAs have the potential to give visibility to hearing loss and relieve some of the stigma surrounding wearable hearing devices. More research is needed, but this increased visibility could positively impact the transition into CIs from patients who would have never considered a CI before. Additionally, amplification use prior to implantation has a positive effect on CI outcomes (84,86,97). Increasing access to amplification benefits the hearing healthcare community at large.

Despite the founding of the American Cochlear Implant Alliance, there is still a need for CI advocacy and awareness campaigns. Social media has allowed CI users from diverse backgrounds and geographies to connect with other users, but these avenues may be more popular amongst younger generations or those with greater access to technology. One study from this review used an awareness campaign to increase CI awareness with suboptimal results (33). Their findings suggested that although individuals were accessing CI manufacturer websites for information, they may not be engaging or reading this information as questions pertaining to HAs and CIs remained unchanged from baseline. CI information available on the internet may not be at an appropriate reading level (60,70). The use of social media to launch an awareness campaign could be more impactful and is something that could potentially be used to address this large barrier globally.

In general, better educational materials, whether through the internet or through healthcare provider networks, may be one way to overcome professional and organizational obstacles. Developing broadly applicable materials that are appropriate across international borders could help educate at the societal level and overcome the obstacle of country-specific implantation criteria. Additionally, having networks of existing CI recipients can help educate and positively influence decision-making in pursuing a CI.

Recently, CI visibility has increased in film and television, which may also increase awareness among the general population. However, this could potentially result in mixed attitudes and beliefs. For example, Sound of Metal (2019) portrayed CIs in a negative light by giving inaccurate information that the device and surgery is not covered by insurance, and the main character ultimately decided not to use their CI possibly due to sound quality. In contrast, Toy Story 4 (2019) displayed the pediatric utilization of CIs in a positive light. In A Quiet Place (2018), the use of a CI in combination with sign language brings to light the utility of CIs in different linguistic and cultural settings. The visibility that we see in entertainment media today may not have been intended to increase awareness but could be looked at as a step in the right direction. Another undoubtedly impactful project was the release of the American Girl doll (2020) who is a CI recipient. This may have a positive impact on future generations though its impact on adult CI utilization is unclear.

The largest research gap identified in this review is the disparity in minority CI utilization. In the US, this population may consist of African Americans, Hispanics, Asians, or anyone of non-Caucasian descent. However, due to the immense size of the US and variability across regions, minority health behavior may present itself differently depending on the area (98). Even less is known about rural and low SES populations which may or may not overlap with racial minority profiles. Rural and remote communities suffer from limited health resources (99). Globally, minority populations may also consist of indigenous persons, cultural immigrants, religious minorities, and the LGBTQIA+ community. It is unknown how these populations are being served but collecting thorough sociocultural data during patient intake may help clinicians to address structural, interpersonal, and individual barriers (100,101). Incorporating this data into our research studies can help gain a better understanding of these unique populations and promote culturally competent care overall.

### Strengths and Limitations

Strengths of this review were that PRISMA-ScR guidelines were followed, the search strategy was expansive and conducted with the help of a university librarian, and the data extraction and screening were done by two independent reviewers. One potential weakness is that some articles included in this review might be deemed as loosely related to barriers to CI uptake, specifically articles related to CI eligibility or utilization (4,21,46). Because of the overarching nature of the ecological framework used and these articles’ ability to capture policy and structural barriers, the articles were included. Other weaknesses include the small sample size in some of the prospective studies as well as the poor methodological validation of tools used in qualitative studies. The prevalence, relative contribution, and intersectionality of individual barriers to cochlear implantation in adults also remain unclear.

### Conclusion

Understanding the body of work surrounding barriers to CI uptake in adults is an important step to providing good clinical care and increasing access and utilization of CIs. The barriers identified in this scoping review fit into an ecological framework that may be helpful when designing targeted interventions to overcome such barriers.

## Supporting information

Mesh Search String

## Data Availability

All data produced in the present work are contained in the manuscript.

