## Supplementary material for "Barriers to Cochlear Implant Uptake in Adults: A Scoping Review": Mesh Search String

**Supplementary Material: MESH Search Terms for PubMed**

("Cochlear Implants"[Mesh] OR "Cochlear Implantation"[Mesh] OR cochlea implant*[tiab] OR "cochlea prostheses"[tiab] OR "cochlea prosthesis"[tiab] OR cochlear implant*[tiab] OR cochlear prostheses[tiab] OR cochlear prosthesis[tiab] OR cochlear prosthetic[tiab] OR cochlear prosthetics[tiab] OR implanted cochlea[tiab] OR implanted cochlear[tiab]) AND ("Black or African American"[Mesh] OR "Black People"[Mesh] OR "Decision Making, Shared"[Mesh] OR "Employment"[Mesh] OR "Ethnic and Racial Minorities"[Mesh] OR "Health Disparate, Minority and Vulnerable Populations"[Mesh] OR "Health Equity"[Mesh] OR "Health Inequities"[Mesh] OR "Health Services Accessibility"[Mesh] OR "Health Status Disparities"[Mesh] OR "Healthcare Disparities"[Mesh] OR "Information Literacy"[Mesh] OR "Insurance"[Mesh] OR "Minority Health"[Mesh] OR "Minority Groups"[Mesh] OR "Patient Acceptance of Health Care"[Mesh:noexp] OR "Patient Participation"[Mesh] OR "Race Factors"[Mesh] OR "Racial Groups"[Mesh] OR "Racism"[Mesh] OR "Referral and Consultation"[Mesh] OR "Social Determinants of Health"[Mesh] OR "Social Discrimination"[Mesh] OR "Social Status"[Mesh] OR "Sociodemographic Factors"[Mesh] OR "Socioeconomic Factors"[Mesh] OR "Vulnerable Populations"[Mesh] OR access to care[tiab] OR "access to CI"[tiab] OR access to cochlear[tiab] OR access to health[tiab] OR access to healthcare[tiab] OR "access to implant*"[tiab] OR access to medical[tiab] OR access to resources[tiab] OR access to service[tiab] OR access to services[tiab] OR access to therap*[tiab] OR access to treatment[tiab] OR access to treatments[tiab] OR accessibility of care[tiab] OR accessibility of health[tiab] OR accessibility of healthcare[tiab] OR accessibility of medical[tiab] OR accessibility of treatment[tiab] OR accessible care[tiab] OR accessible health[tiab] OR accessible healthcare[tiab] OR accessible medical[tiab] OR accessible therap*[tiab] OR accessible treatment[tiab] OR accessible treatments[tiab] OR African American[tiab] OR African Americans[tiab] OR African ancestries[tiab] OR African ancestry[tiab] OR African continental ancestry[tiab] OR Afro-American[tiab] OR Afro-Americans[tiab] OR ancestry group[tiab] OR ancestry groups[tiab] OR antiracism[tiab] OR availability of care[tiab] OR availability of health[tiab] OR availability of healthcare[tiab] OR availability of medical[tiab] OR availability of treatment[tiab] OR available care[tiab] OR available health[tiab] OR available healthcare[tiab] OR available medical[tiab] OR available therap*[tiab] OR available treatment[tiab] OR available treatments[tiab] OR barrier[tiab] OR barriers[tiab] OR Black[tiab] OR Blacks[tiab] OR care accept*[tiab] OR care access[tiab] OR care accessibility[tiab] OR care availab*[tiab] OR care deliver*[tiab] OR care disparit*[tiab] OR care eligib*[tiab] OR care right[tiab] OR care rights[tiab] OR care seeking behav*[tiab] OR care use[tiab] OR care utilisation[tiab] OR care utilization[tiab] OR CI utilization[tiab] OR consultation[tiab] OR consultations[tiab] OR determining health[tiab] OR determining factor[tiab] OR determining factors[tiab] OR disadvantaged[tiab] OR disparate group[tiab] OR disparate groups[tiab] OR disparate population[tiab] OR disparate populations[tiab] OR disparities in care[tiab] OR disparities in health[tiab] OR disparities in healthcare[tiab] OR disparities in treatment[tiab] OR disparity in health[tiab] OR disparity in healthcare[tiab] OR economic characteristics[tiab] OR economic class[tiab] OR economic classes[tiab] OR economic condition[tiab] OR economic conditions[tiab] OR economic connectedness[tiab] OR economic dependence[tiab] OR economic factor[tiab] OR economic factors[tiab] OR economic independence[tiab] OR economic policies[tiab] OR economic policy[tiab] OR economic standing[tiab] OR economic status[tiab] OR economic statuses[tiab] OR econsultation[tiab] OR econsultations[tiab] OR education achieve*[tiab] OR education attain*[tiab] OR education index[tiab] OR education level[tiab] OR education levels[tiab] OR education status[tiab] OR educational achieve*[tiab] OR educational attain*[tiab] OR educational index[tiab] OR educational level[tiab] OR educational levels[tiab] OR educational status[tiab] OR ehealth literacy[tiab] OR ehealth literate[tiab] OR employment[tiab] OR employments[tiab] OR equalities[tiab] OR equality[tiab] OR equit*[tiab] OR ethnic*[tiab] OR ethno-linguistic[tiab] OR ethnolinguistic[tiab] OR facilitating factor[tiab] OR facilitating factors[tiab] OR facilitator[tiab] OR facilitators[tiab] OR family characteristics[tiab] OR family demograph*[tiab] OR family dynamic[tiab] OR family dynamics[tiab] OR family life[tiab] OR family status[tiab] OR family statuses[tiab] OR financial burden[tiab] OR financial burdens[tiab] OR financial condition[tiab] OR financial conditions[tiab] OR financial constraint[tiab] OR financial constraints[tiab] OR financial dependence[tiab] OR financial implication[tiab] OR financial implications[tiab] OR financial independence[tiab] OR financial status[tiab] OR financial statuses[tiab] OR financially dependent[tiab] OR financially independent[tiab] OR gate keep*[tiab] OR gatekeep*[tiab] OR geographic access[tiab] OR geographic accessibility[tiab] OR geographic availab*[tiab] OR geographic burden[tiab] OR geographic disparit*[tiab] OR geographical access[tiab] OR geographical accessibility[tiab] OR geographical availab*[tiab] OR geographical burden[tiab] OR geographical disparit*[tiab] OR group discriminat*[tiab] OR group disparit*[tiab] OR groups discriminat*[tiab] OR health determinant[tiab] OR health determinants[tiab] OR health discriminat*[tiab] OR health disparit*[tiab] OR health literacy[tiab] OR health literate[tiab] OR health right[tiab] OR health rights[tiab] OR health status[tiab] OR health utilisation[tiab] OR health utilization[tiab] OR healthcare accept*[tiab] OR healthcare access[tiab] OR healthcare accessibility[tiab] OR healthcare availab*[tiab] OR healthcare disparit*[tiab] OR healthcare eligib*[tiab] OR healthcare right[tiab] OR healthcare rights[tiab] OR healthcare seeking behav*[tiab] OR healthcare use[tiab] OR healthcare utilisation[tiab] OR healthcare utilization[tiab] OR housing eviction[tiab] OR housing evictions[tiab] OR housing insecurities[tiab] OR housing insecurity[tiab] OR housing instability[tiab] OR implant utilization[tiab] OR implantation utilization[tiab] OR income[tiab] OR indigence[tiab] OR indigency[tiab] OR indigent health[tiab] OR indigent healthcare[tiab] OR inequal*[tiab] OR inequit*[tiab] OR information literacy[tiab] OR information literate[tiab] OR insecure housing[tiab] OR insecure residen*[tiab] OR insurance[tiab] OR level of education*[tiab] OR limited care[tiab] OR limited health[tiab] OR limited healthcare[tiab] OR limited resource[tiab] OR limited resources[tiab] OR limited service[tiab] OR limited services[tiab] OR living condition[tiab] OR living conditions[tiab] OR living standard[tiab] OR living standards[tiab] OR low utilisation[tiab] OR low utilization[tiab] OR lower class[tiab] OR lower classes[tiab] OR lower utilisation[tiab] OR lower utilization[tiab] OR marital status[tiab] OR marital statuses[tiab] OR marriage status[tiab] OR medical access[tiab] OR medical accessibility[tiab] OR medical availab*[tiab] OR medical eligib*[tiab] OR medical right[tiab] OR medical rights[tiab] OR medication access[tiab] OR medication accessibility[tiab] OR medication availab*[tiab] OR medication eligib*[tiab] OR medicine access[tiab] OR medicine accessibility[tiab] OR medicine availab*[tiab] OR middle class[tiab] OR middle classes[tiab] OR minorities[tiab] OR minority[tiab] OR Negro*[tiab] OR non-employed[tiab] OR nonemployed[tiab] OR occupation status[tiab] OR occupation statuses[tiab] OR occupational status[tiab] OR occupational statuses[tiab] OR patient accept*[tiab] OR patient activation[tiab] OR patient attitude[tiab] OR patient attitudes[tiab] OR patient empowerment[tiab] OR patient engagement[tiab] OR patient involvement[tiab] OR patient nonaccept*[tiab] OR patient participation[tiab] OR people of color[tiab] OR person of color[tiab] OR population discriminat*[tiab] OR population disparit*[tiab] OR poverty[tiab] OR race based[tiab] OR race bias[tiab] OR race biases[tiab] OR race discriminat*[tiab] OR race disparit*[tiab] OR race factor[tiab] OR race factors[tiab] OR race motivated[tiab] OR race prejudice[tiab] OR race related[tiab] OR racial bias[tiab] OR racial biases[tiab] OR racial discriminat*[tiab] OR racial disparit*[tiab] OR racial factor[tiab] OR racial factors[tiab] OR racial prejudice[tiab] OR racial prejudices[tiab] OR racially based[tiab] OR racially biased[tiab] OR racially motivated[tiab] OR racism[tiab] OR referral[tiab] OR referrals[tiab] OR residential eviction[tiab] OR residential evictions[tiab] OR residential instability[tiab] OR resource access[tiab] OR resource accessibility[tiab] OR resource availab*[tiab] OR resource deliver*[tiab] OR resource disparit*[tiab] OR resource eligib*[tiab] OR resource use[tiab] OR resource utilisation[tiab] OR resource utilization[tiab] OR resources access[tiab] OR resources accessibility[tiab] OR resources availab*[tiab] OR resources utilisation[tiab] OR resources utilization[tiab] OR right to care[tiab] OR right to health[tiab] OR right to healthcare[tiab] OR right to medical[tiab] OR right to treatment[tiab] OR right to treatments[tiab] OR rights to health[tiab] OR rights to healthcare[tiab] OR second opinion[tiab] OR second opinions[tiab] OR sensitive group[tiab] OR sensitive groups[tiab] OR sensitive people[tiab] OR sensitive population[tiab] OR sensitive populations[tiab] OR service access[tiab] OR service accessibility[tiab] OR service availab*[tiab] OR service deliver*[tiab] OR service disparit*[tiab] OR service eligib*[tiab] OR service right[tiab] OR service use[tiab] OR service utilisation[tiab] OR service utilization[tiab] OR services access[tiab] OR services accessibility[tiab] OR services availab*[tiab] OR services deliver*[tiab] OR services disparit*[tiab] OR services eligib*[tiab] OR services right[tiab] OR services use[tiab] OR services utilisation[tiab] OR services utilization[tiab] OR social attribute[tiab] OR social attributes[tiab] OR social class[tiab] OR social classes[tiab] OR social characteristics[tiab] OR social condition[tiab] OR social conditions[tiab] OR social connectedness[tiab] OR social cultur*[tiab] OR social demograph*[tiab] OR social dependence[tiab] OR social determinant[tiab] OR social determinants[tiab] OR social discriminat*[tiab] OR social disparit*[tiab] OR social econom*[tiab] OR social factor[tiab] OR social factors[tiab] OR social health[tiab] OR social healthcare[tiab] OR social identities[tiab] OR social identity[tiab] OR social independence[tiab] OR social policies[tiab] OR social policy[tiab] OR social politics[tiab] OR social standing[tiab] OR social status[tiab] OR social statuses[tiab] OR socially connected[tiab] OR socially dependent[tiab] OR socially independent[tiab] OR socio-cultur*[tiab] OR socio-demograph*[tiab] OR socio-econom*[tiab] OR sociocultur*[tiab] OR sociodemograph*[tiab] OR socioeconom*[tiab] OR standard living[tiab] OR standard of living[tiab] OR structural determinant[tiab] OR structural determinants[tiab] OR teleconsultation[tiab] OR teleconsultations[tiab] OR travel burden[tiab] OR treatment access[tiab] OR treatment accessibility[tiab] OR treatment availab*[tiab] OR treatment discriminat*[tiab] OR treatment disparit*[tiab] OR treatment eligib*[tiab] OR treatment right[tiab] OR treatment rights[tiab] OR treatment use[tiab] OR treatment utilisation[tiab] OR treatment utilization[tiab] OR un-employed[tiab] OR under-represented[tiab] OR under-resourced[tiab] OR under-served[tiab] OR under-utilis*[tiab] OR under-utiliz*[tiab] OR underrepresented[tiab] OR underresourced[tiab] OR underserved[tiab] OR underutilis*[tiab] OR underutiliz*[tiab] OR unemployed[tiab] OR unemployment[tiab] OR unemployments[tiab] OR unmet care[tiab] OR unmet health[tiab] OR unmet healthcare[tiab] OR unmet medical[tiab] OR unstable econom*[tiab] OR unstable housing[tiab] OR unstable residen*[tiab] OR upper class[tiab] OR upper classes[tiab] OR uptake[tiab] OR use of care[tiab] OR use of health[tiab] OR use of healthcare[tiab] OR use of medical[tiab] OR use of service[tiab] OR use of services[tiab] OR use of treatment[tiab] OR "utilisation of cochlear"[tiab] OR utilisation of health[tiab] OR utilisation of healthcare[tiab] OR utilisation of medical[tiab] OR utilisation rate[tiab] OR utilisation rates[tiab] OR utilization of care[tiab] OR "utilization of CI"[tiab] OR "utilization of cochlear"[tiab] OR utilization of health[tiab] OR utilization of healthcare[tiab] OR utilization of medical[tiab] OR utilization rate[tiab] OR utilization rates[tiab] OR vulnerable group[tiab] OR vulnerable groups[tiab] OR vulnerable people[tiab] OR vulnerable person[tiab] OR vulnerable persons[tiab] OR vulnerable population[tiab] OR vulnerable populations[tiab] OR (("Patients"[Mesh] OR candidacy[tiab] OR candidate*[tiab] OR informed[tiab] OR patient*[tiab] OR recipient*[tiab] OR shared[tiab]) AND ("Decision Making"[Mesh] OR decision*[tiab] OR defer*[tiab] OR hesitan*[tiab] OR refus*[tiab] OR reluctan*[tiab]))) NOT (("Adolescent"[Mesh] OR "Child"[Mesh] OR "Infant"[Mesh]) NOT "Adult"[Mesh])
